# A spatio-temporal framework for modelling wastewater concentration during the COVID-19 pandemic

**DOI:** 10.1101/2022.10.14.22281081

**Authors:** Guangquan Li, Hubert Denise, Peter Diggle, Jasmine Grimsley, Chris Holmes, Daniel James, Radka Jersakova, Callum Mole, George Nicholson, Camila Rangel Smith, Sylvia Richardson, William Rowe, Barry Rowlingson, Fatemeh Torabi, Matthew J. Wade, Marta Blangiardo

## Abstract

The potential utility of wastewater-based epidemiology as an early warning tool has been explored widely across the globe during the current COVID-19 pandemic. Methods to detect the presence of SARS-CoV-2 RNA in wastewater were developed early in the pandemic, and extensive work has been conducted to evaluate the relationship between viral concentration and COVID-19 case numbers at the catchment areas of sewage treatment works (STWs) over time. However, no attempt has been made to develop a model that predicts wastewater concentration at fine spatio-temporal resolutions covering an entire country, a necessary step towards using wastewater monitoring for the early detection of local outbreaks.

We consider weekly averages of flow-normalised viral concentration, reported as the number of SARS-CoV-2 N1 gene copies per litre (gc/L) of wastewater available at 303 STWs over the period between 1 June 2021 and 30 March 2022. We specify a spatially continuous statistical model that quantifies the relationship between weekly viral concentration and a collection of covariates covering socio-demographics, land cover and virus-associated genomic characteristics at STW catchment areas while accounting for spatial and temporal correlation.

We evaluate the model’s predictive performance at the catchment level through 10-fold cross-validation. We predict the weekly viral concentration at the population-weighted centroid of the 32,844 lower super output areas (LSOAs) in England, then aggregate these LSOA predictions to the Lower Tier Local Authority level (LTLA), a geography that is more relevant to public health policy-making. We also use the model outputs to quantify the probability of local changes of direction (increases or decreases) in viral concentration over short periods (e.g. two consecutive weeks).

The proposed statistical framework is able to predict SARS-CoV-2 viral concentration in wastewater at high spatio-temporal resolution across England. Additionally, the probabilistic quantification of local changes can be used as an early warning tool for public health surveillance.

## 1. Introduction

Wastewater-based epidemiology (WBE) is defined as a collection of tools and methods for surveillance and monitoring disease outbreaks using biochemical analysis of wastewater samples as the primary outcome measure. The first use of WBE was to track illicit drug use (see for instance Daughton (2001) as the first published paper on the topic and Huizer et al. (2021) for a recent review of the field). Over the years, WBE has been successfully used in polio eradication (Asghar et al., 2014; Hovi et al., 2012) and for retrospective prediction of several disease outbreaks, such as noroviruses and hepatitis A (Hellmér et al., 2014), especially in resource-limited settings. During the COVID-19 pandemic, WBE has been recognised as an economically efficient approach for disease surveillance (Manuel et al., 2022) and methods to detect the presence of SARS-CoV-2 RNA in wastewater have been developed in a number of countries (Tlhagale et al., 2022). In addition to its relatively low cost, an attraction of working with wastewater is that it avoids the selection bias that is an inherent feature of other widely used pandemic metrics such as community testing. However, viral load can be affected by several factors other than local prevalence of COVID-19, including meteorology, type of sewage system, and population characteristics; for example, children shed virus at a lower rate than adults (Wade et al., 2022). Additionally, the relationship between wastewater viral load and COVID-19 prevalence is susceptible to changes in the nature of the epidemic over time, such as vaccination rates or the shedding properties and epidemiology of different variants (Wu et al., 2020; Nattino et al., 2022).

Several studies have evaluated the relationship between viral load in wastewater and prevalence of COVID-19 disease. In particular, Shah et al. (2022) conducted a systematic review of wastewater surveillance methods for monitoring the COVID-19 pandemic. They focused on the period between 1 January 2020 and 31 July 2021, and identified 84 studies spanning 34 countries that reported a potential relationship between viral concentration in wastewater and COVID-19 cases in the community. In England, Hillary et al. (2021) quantified concentration of SARS-CoV-2 RNA from six sewage treatment works (STWs) in large urban centres during the first wave of the pandemic (March-July 2020) and reported a correlation with number of COVID-19 cases. More specifically, they found that a decline in the wastewater virus concentration preceded by two to four days the reduction in community cases after lockdown measures were implemented. More recently, Morvan et al. (2022) analysed data from 45 STWs across England between July 2020 and March 2021. Using a multilevel modelling approach they reported that wastewater samples can be used successfully to predict COVID-19 prevalence, with a lead time of four to five days. Proverbio et al. (2022) proposed a mechanistic approach to integrate wastewater data and COVID-19 case numbers across different countries using an extended Kalman filter (Durbin and Koopman, 2012, Chapter 10), and showed how this can be used for early detection of outbreaks. Srinivas et al. (2021) used Bayesian networks to evaluate the best geographical locations and population characteristics for using wastewater to detect regions of outbreaks over 13 US states.

Scientific contributions to date have focused on STW sites and the associated catchment areas where the measurements of RNA from wastewater are obtained. No attempt has been made to develop a spatially resolved model to predict wastewater concentration over a spatially continuous domain, required for the use of wastewater as a tool for the early detection of local outbreaks. Geostatistical methods (Diggle and Ribeiro, 2007; Diggle et al., 1998) are naturally suited to this task, as they allow the combination of observations on the outcome variable of interest at a fixed set of point locations with a set of predictors available at fine-scale spatial resolution throughout the area of interest. They account for a combination of covariate effects and residual spatial and temporal structure to deliver predictions on the outcome at any point in space. We specify a Bayesian geostatistical model which quantifies the relationship between weekly viral concentration at STW catchment areas and covariates (socio-demographics, land cover and virus genomic properties) while accounting for spatial and temporal correlation. We then use the model to predict weekly viral concentration together with the associated predictive uncertainty at the population weighted centroids of Lower Super Output Areas (LSOAs), a set of small geographical areas used in census reporting in the UK. These LSOA predictions can be combined to produce predictions at coarser geographical scales. We present results at the Lower Tier Local Authoritie (LTLA) level, larger administrative areas than LSOAs, as an example of a geographical scale that is relevant for public health policies. In addition, the probabilistic output from this flexible modelling framework can be used to make a variety of predictive inferences, for example to detect areas where the level of viral concentration in wastewater exceeds a pre-defined threshold or where the temporal pattern shows increases over consecutive weeks.

## 2. Methods

### 2.1. Study area and Data

SARS-CoV-2 viral concentrations were obtained through reverse transcriptase quantitative polymerase chain reaction (RT-qPCR) analysis of wastewater samples, as described in Hillary et al. (2021). Three to four weekly samples were taken at each sewer network sites (serving local areas) and STWs (serving cities or towns) by the Environmental Monitoring for Health Protection (EMHP) wastewater surveillance programme (Wade et al., 2020), formerly part of the Joint Biosecurity Centre, now UK Health Security Agency (UKHSA). In the present study we consider only the 303 STWs for which the data were publicly available from EMHP. The locations of the STWs and their catchment areas are presented in Figure 1 (A).

**Figure 1:**
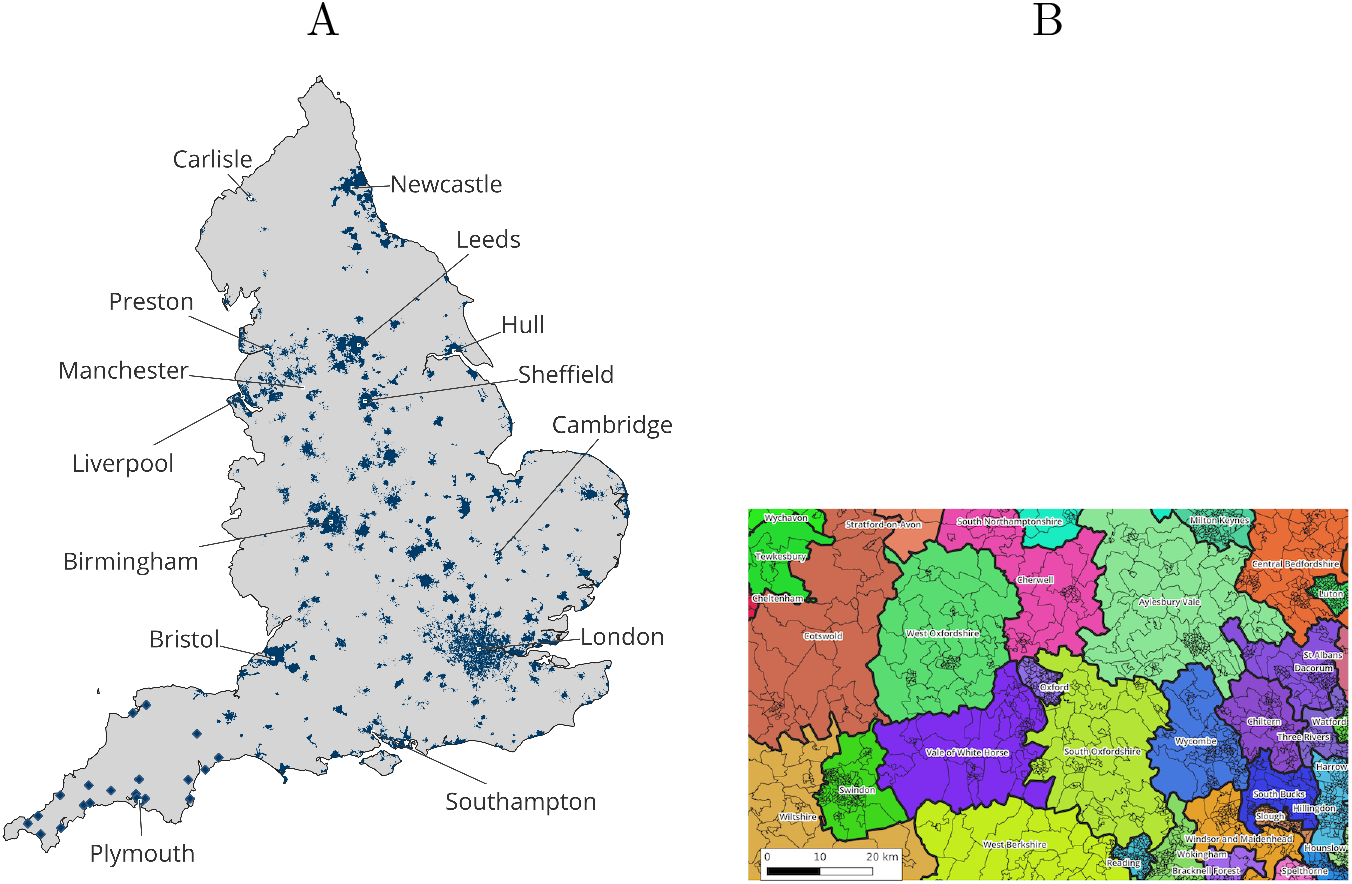
(A): Locations of 303 STW catchment areas in England (blue); (B): LTLA and LSOA boundaries for a representative part of southern England.

The data that we use in our analysis consist of the weekly average of the three/four flow-normalised viral concentration measurements, reported as the number of SARS-CoV-2 N1 gene copies per litre of wastewater (gc/L), at each STW over the period from 1 June 2021 to 30 March 2022.

We include the following covariates to inform the spatio-temporal variability of viral concentration in wastewater:

- Index of Multiple Deprivation(IMD, 2019). This composite index is a weighted average of seven aspects of socio-economic deprivation: Income; Employment; Health Deprivation and Disability; Education, Skills Training; Crime; Barriers to Housing and Services; Living Environment.
- Black, Asian and Minority Ethnic (BAME) proportion in each area as reported in the 2011 Census.
- Land cover, calculated by intersecting boundaries with the latest (2018) Corine Land Cover data set and computing the total fraction of area in urban, vegetation, industrial, and “all other” classes. The data is on a 1km grid and available from https://land.copernicus.eu/pan-european/corine-land-cover and © European Union, Copernicus Land Monitoring Service 2018, European Environment Agency (EEA).
- Population density, estimated by the Office for National Statistics (ONS) in 2019.
- Age structure, defined as the percentages of population younger than 16 years and older than 75 years, estimated by the ONS in 2019.
- Wastewater genomic data have been collected by UKHSA and shown generally high correlation with clinical cases over time, suggesting that they could be used as predictors in our model. We consider the percentage coverage of the SARS-CoV-2 genome in each sample, as obtained from position read file, and the Single-Nucleotide Polymorphism (SNP) data. These two measures are included as time-varying covariates at the national level. For both covariates, there were no measurements taken over the week commencing 28 March, 2022, the last week of the study period. We impute the missing value of that week by using the previous week’s value. For a majority of the 43 weeks, the genomic measurements were taken at over 80% of the 303 STW sites but there are still some weeks where measurements were only taken at a small number of sites. A rolling average over three weeks on either side is therefore applied to obtain reliable national averages and the resulting averages entered the model.

If not explicitly stated above, the covariates are available at LSOA level. We map STW catchment areas to LSOAs by comparing their respective boundaries and aligning the covariates to STW catchment areas as follows: (i) for population density we sum over the LSOAs within the geographical boundaries of the catchment; (ii) for IMD, BAME and age structure we calculate the population-weighted average over the corresponding LSOAs. As land cover is available at grid level we average the grid values covering each LSOA and catchment areas to obtain the variable at each of these geographical scales. The LSOA-to-catchment mapping is carried out via the lookup table from (Hoffmann et al., 2022) which was created based on the wastewater catchment area data provided by the sewerage service providers in Great Britain. This look-up table covers all STWs in England except the 21 STWs in the South West, the locations with a diamond shape in Figure 1A. For these 21 STWs, we approximated their catchments using circles that are centered at the STW locations and include the centroids of at least 10 LSOAs.

Our approach enables prediction SARS-CoV-2 viral concentration at the highest spatial resolution for which all of the relevant covariate are available, while fully accounting for spatial variability. These predictions can then be aggregated to other geographical scales that might be more relevant for public health policies. With this in mind, we predict concentration at LSOA level in England and then aggregate these to LTLA level; see Figure 1(B) for an example of LSOA and LTLA boundaries in a representative part of England. LSOAs are intended to include approximately 1,500 individuals and therefore cover smaller areas in densely populated areas. When aggregating from LSOA to LTLA level we used the 2019 LSOA population estimates as weights.

### 2.2. Statistical model

The log-transformed number of gene copies per liter (henceforth *concentration*) at catchment area *i* = 1, …, 303 and week *t* = 1, …, 44 is modelled as:

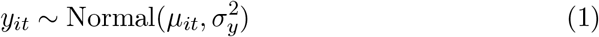

where 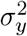 is the measurement error variance. For the latent mean concentration *μ*_*it*_ we specify a linear model:

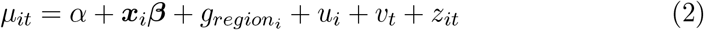

where *α* is the average concentration across the study area and ***x***_*it*_ = {*x*_1*it*_, *x*_2*it*_, …, *x*_*mit*_} is the vector of *m* = 8 covariates as specified above. The 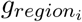 term is a regional-level random effect with *region*_*i*_ indicating the region in which STW *i* is located. The regional random effects are modelled using an exchangeable prior, 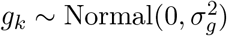 with *k* = 1, …, 9 over the nine regions of England. The *u*_*i*_ term is a catchment-level spatial random effect modelled using an exchangeable prior, 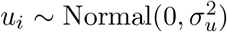. The *v*_*t*_ term is a temporal random effect, which we model as a first-order random walk, 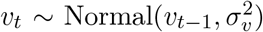. Finally, we allow for additional flexibility by including a spatio-temporal interaction component, *z*_*it*_, to capture local departures from the global spatial and temporal patterns. For this, we specify a temporal autoregressive structure with spatially correlated innovations, similar to Cameletti et al. (2011):

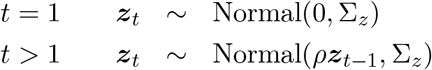

with *ρ* as the temporal autoregressive coefficient. The ∑_*z*_ matrix represents the spatial structure and is defined using a Matèrn covariance function:

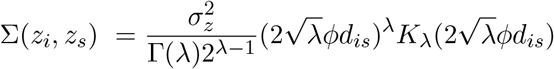

where Γ(*λ*) is the gamma function, *K*_*λ*_ is the modified Bessel function of second kind with order *λ* and *d*_*is*_ is the distance between the *i*-th and *s*-th sites. The parameter *ϕ >* 0 controls the rate of decay of the correlation as the distance *d* increases, while *λ >* 0 controls the smoothness of the ***z*** random field. As it is common practice to avoid issues with identifiability, we fix the value *λ* = 1 to correspond to a mean-square differentiable spatial process (Diggle and Ribeiro, 2007; Whittle, 1954).

#### 2.2.1. Priors and implementation

We complete the Bayesian model specification by assigning the following priors. On the intercept *α* and the fixed effects ***β*** in Eq.(2) we assign independent Normal distributions centered on 0 with variance equal to 10^3^. All covariates were standardised so this specification corresponds to a minimally informative prior on the covariate effect. Following Simpson et al. (2017), on the random effect variances and parameters associated with the spatio-temporal interaction component we specify *Penalised Complexity* (PC) priors that are defined via probability statements. Specifically, the PC prior on 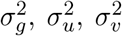 and 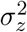 is specified based on *Pr*(*σ >* 10) = 0.05. Given the log-scale of the viral concentration, this specification is weakly informative, assuming that the standard deviations are highly likely to be between 0 and 10, with only a small probability being greater than 10. For the Matèrn covariance function, we specify a PC prior on the correlation range *r*, which we define <u>as</u> the d<u>i</u>stance at which the correlation is approximately 0.1, hence 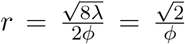. We specify the PC prior such that *Pr*(*r <* 10) = 0.05, highly likely that the range is greater than 10 km. On the autoregressive parameter *ρ*, the PC prior is defined such that *Pr*(|*ρ*| *>* 0.1) = 0.9. Finally, a weakly informative Gamma prior Gamma(1, 0.00005) is assigned to the error precision, 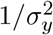.

The inferential task is to estimate the joint posterior distribution of the regression coefficients, *α*, ***β***, the regional, space, time and space-time random effects ***g, u, v, z*** and the parameters 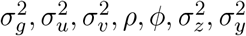:

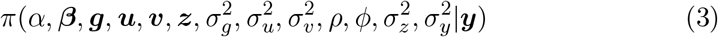

We implement the model in R-INLA (Rue et al., 2009), an inferential method based on Integrated Nested Laplace Approximations (INLA). INLA provides fast approximation of the posterior and predictive distributions by exploiting conditional independence on the structure of the model parameters. When the interest is in a continuous spatial domain and the data are available at point locations (as in the current case, where we have the coordinates of the STWs), R-INLA can be coupled with Stochastic Partial Differential Equations (SPDE) (Lindgren et al., 2010). In order to provide fast inference, SPDE discretises the continuous space using weighted basis functions defined at the vertices of a triangulation (mesh) of the study-region. We use the mesh shown in Figure 2. More details on the INLA and SPDE approach are available at Krainski et al. (2018) or Blangiardo and Cameletti (2015).

**Figure 2:**
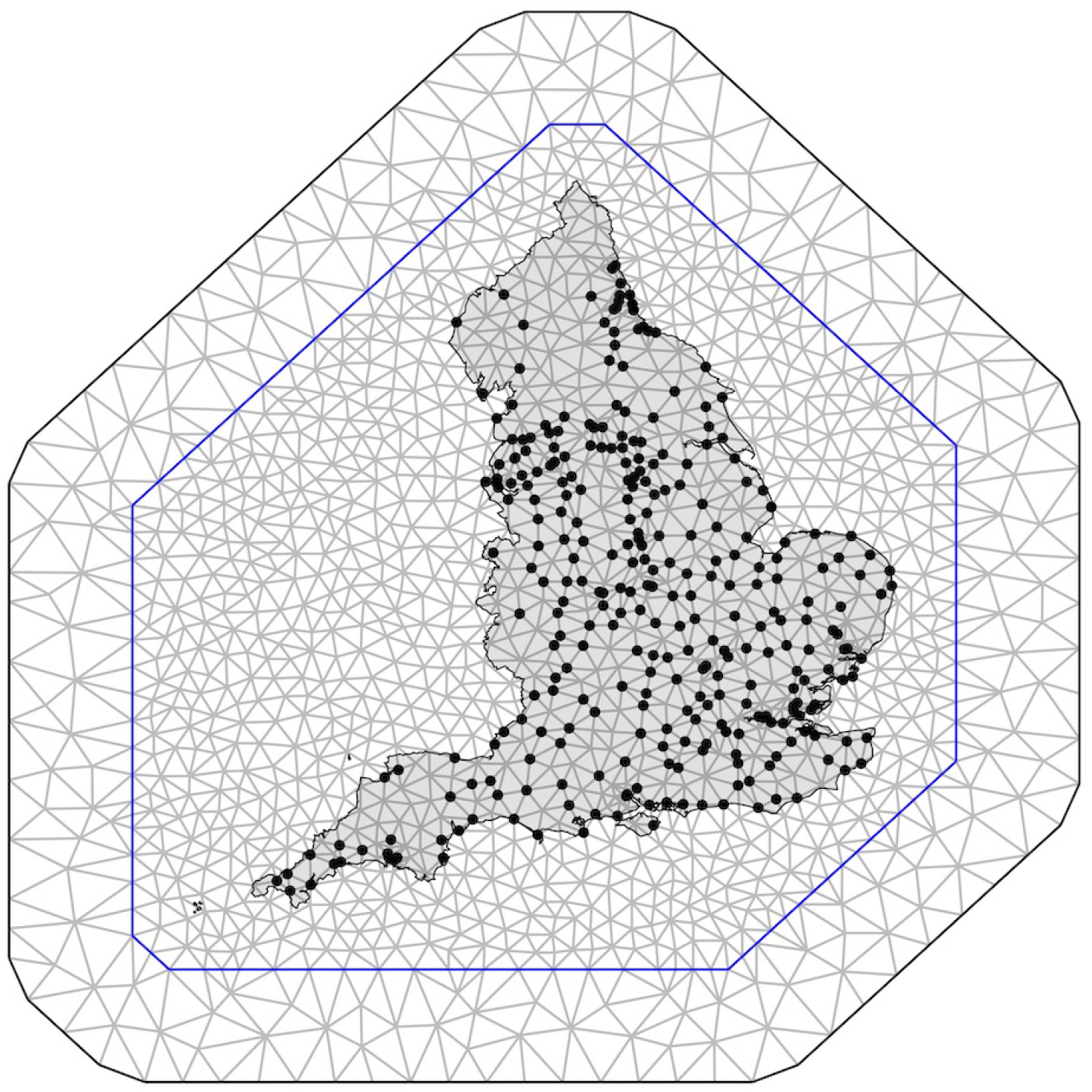
Mesh used for model fitting in INLA with England boundary superimposed. The solid dots represent the locations of the 303 sewage treatment works (STWs) included in the study.

The code and the data to reproduce the results are available at https://github.com/gqlNU/publicWW/.

### 2.3. Predicting viral concentration at a set of prediction locations

A strength of the modelling approach developed here is the ability to use the posterior distribution in Eq.(3) to predict the concentrations at any set of spatial locations in the study region. In particular, we consider the population-weighted centroids of all the 32,844 Lower Super Output Areas (LSOAs) in England, these being the smallest areas for which all the covariates are available.

The weekly concentration of the *j* − *th* LSOA is calculated based on Eq.(2):

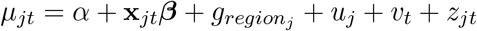

where **x**_*jt*_ denotes the covariate profile of the j − *th* LSOA at time *t* and *region*_*j*_ indicates the region in which this LSOA is located. Given observed data ***y*** we sample ***y***^*t*^, the viral concentrations at the 32,844 LSOA centroids over 44 weeks, from the posterior predictive distribution:

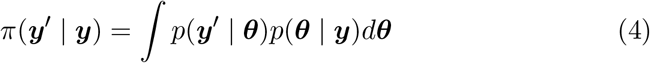

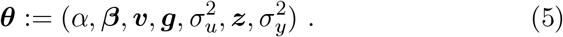

In practice, this involves sampling values of ***θ*** directly from the joint posterior distribution at Eq.(3) followed by the sampling of *u*. For *u*_*j*_, having first sampled 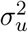 from the joint posterior, we sample *u*_*j*_ via 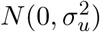. Effectively, *u*_*j*_ adds random noise to the predictions of each LSOA, reflecting the between-site variability estimated from the data, so widening the predictive interval.

The posterior sampling of the space-time interaction component, *z*_*jt*_, at a prediction location is somewhat more involved because its values at non-mesh points are not automatically outputted in the joint posterior. From the joint posterior, we first sample jointly *z*_*bt*_ (*b* = 1, …, *B*; *t* = 1, …, 44), all the terms in the space-time interaction component across all time points and all *B* locations that form part of the SPDE mesh used for model-fitting (see Figure 2). We then project each time slice of the sampled vector ***z***_1:*B,t*_ onto a mesh that extends the fitting mesh to include all the prediction locations, i.e. in our case all LSOA centroids, to form *z*_*jt*_ for *j* = 1, …, 32844, as required to complete the weekly LSOA-level prediction. As space-time interactions are strongly correlated both spatially and temporally, the prediction of this component must be carried out jointly over space and time to account for the space-time dependency.

Finally, the posterior predictive distribution at the LSOA centroids can be combined to return corresponding predictions at coarser geographical scales, as required. Here we consider all the 309 LTLAs in England by averaging the corresponding LSOA predictions, weighted by the population in each LSOA.

### 2.4. Cross-validation

We evaluate the model predictive performance via 10-fold cross validation. We partition all the 303 sites randomly into 10 subgroups, 9 groups of 30 sites and one group of 33. For each of the 10 cross-validation runs, we leave out the viral concentration data in one subgroup in turn, fit the model to the data in the remaining 9 subgroups, then predict the weekly viral concentrations for each of the sites in the left-out group. To assess the agreement between the predicted and observed concentrations at each left-out site we use the following metrics: a) the mean bias and the mean absolute bias with bias defined as the difference between predicted and observed values; b) the root mean square error, defined as the square root of the average of the squared differences between predicted and observed values; and c) the 95% coverage, defined as the percentage of the observed concentration values that lie within the 95% predictive intervals from the model.

### 2.5. Detection

The posterior predictive distribution can be used to detect increases in the wastewater viral concentration. In particular, for each LTLA (*l* = 1, …, *L*) an increase in viral concentration over two consecutive weeks is the event (E):

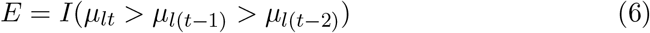

where *I*(·) is the indicator function. A high predictive probability, Prob(*E*) *> c* for a pre-specified threshold *c*, would then trigger a warning of a potential outbreak. This approach has been used for public health surveillance in both high-income (Diggle et al., 2005) and low-to-middle-income countries (Diggle et al., 2007). Selecting the threshold is a key point: lower and higher values increase the false positive and false negative rates, respectively. We follow the recommendation in Richardson et al. (2004) that *c* = 0.8 is a good compromise to minimise the overall false detection rate, whilst recognising that in specific applications it may be preferable to prioritise protection against one or other of the two kinds of detection error.

## 3. Results

### 3.1. Spatial and temporal patterns

Viral concentrations in wastewater varied substantially across England over the observation period between June 1, 2021 and March 30, 2022. Figure 3 (A) presents the overall time trend of log-concentration in England. A sharp increase is visible at the beginning of the study period, with a peak around 10 log(gc/L) in July-August 2021. A second increase is seen around November 2021, with a peak just after Christmas, followed by a final increase in February-March 2022. Figures 3 (B) and (C) visualise the LTLA-level and regional-level variation, respectively. Across the nine regions of England, London shows the highest overall viral concentration level. North West and Yorkshire and The Humber are the two regions with the lowest levels of viral concentration across the study period.

**Figure 3:**
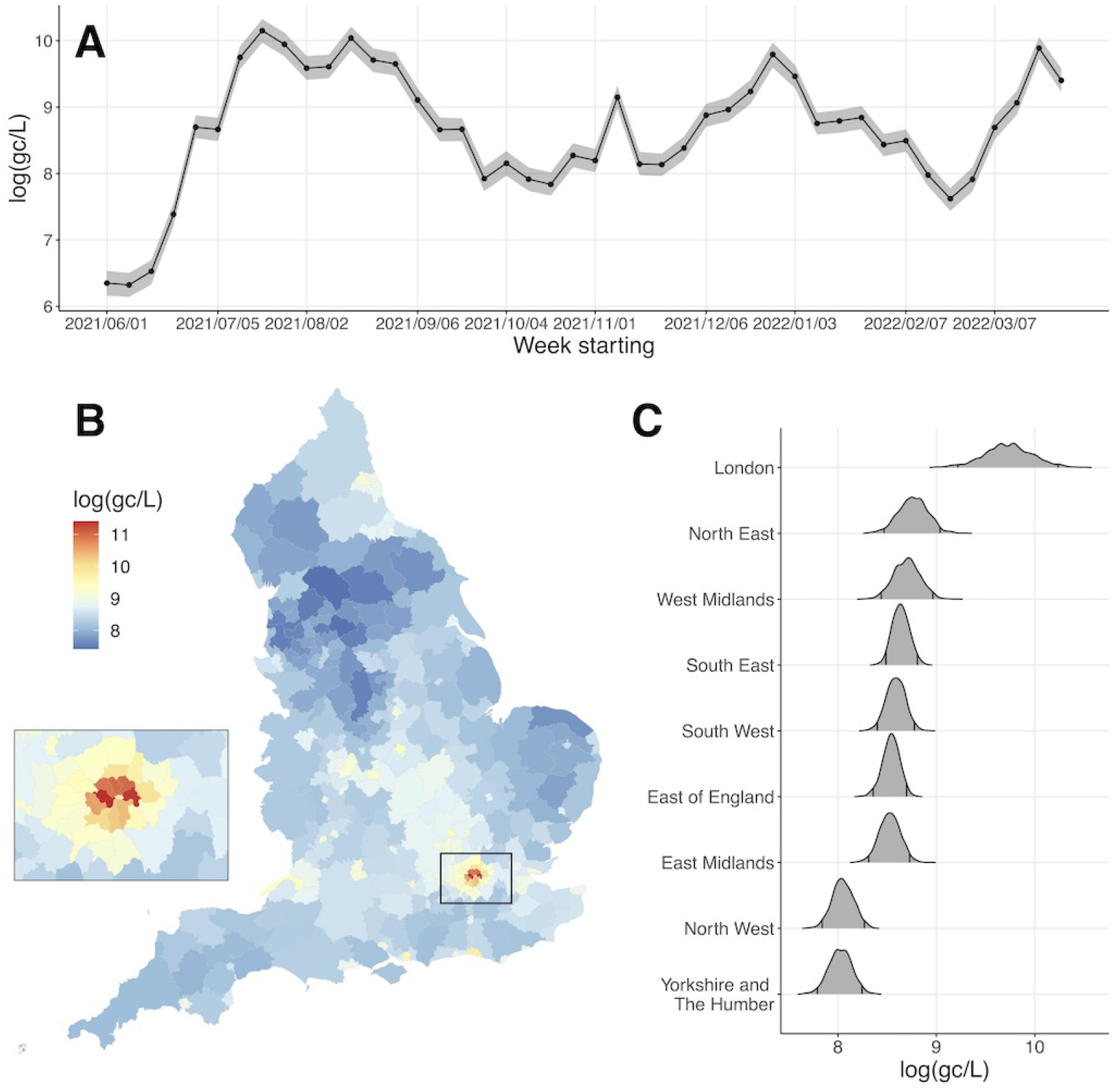
Posterior mean and 95% credible interval of weekly wastewater viral concentration at country level (A); posterior mean at LTLA level (B) and posterior distribution at regional level (C).

### 3.2. Spatio-temporal dynamics

Figure 4 offers further insights into the spatio-temporal dynamics of viral concentration, showing the posterior mean of the predicted log concentration for the first week of the study period (June 1-6, 2021) then one week in every four till March 30, 2022. The rise and fall of viral concentrations at the national level did not happen at the same time across all LTLAs. At the beginning of June 2021 (Figure 4 top, left), low values are estimated across most LTLAs while inner London shows higher values, around 8-9 log(gc/L). By mid-July 2021, an increment is clearly visible across the country, but a high degree of variation is noticeable, with the highest values in pockets of LTLAs in the north, around East Midlands and for the whole of London, whereas parts of East and South West remain relatively low.

**Figure 4:**
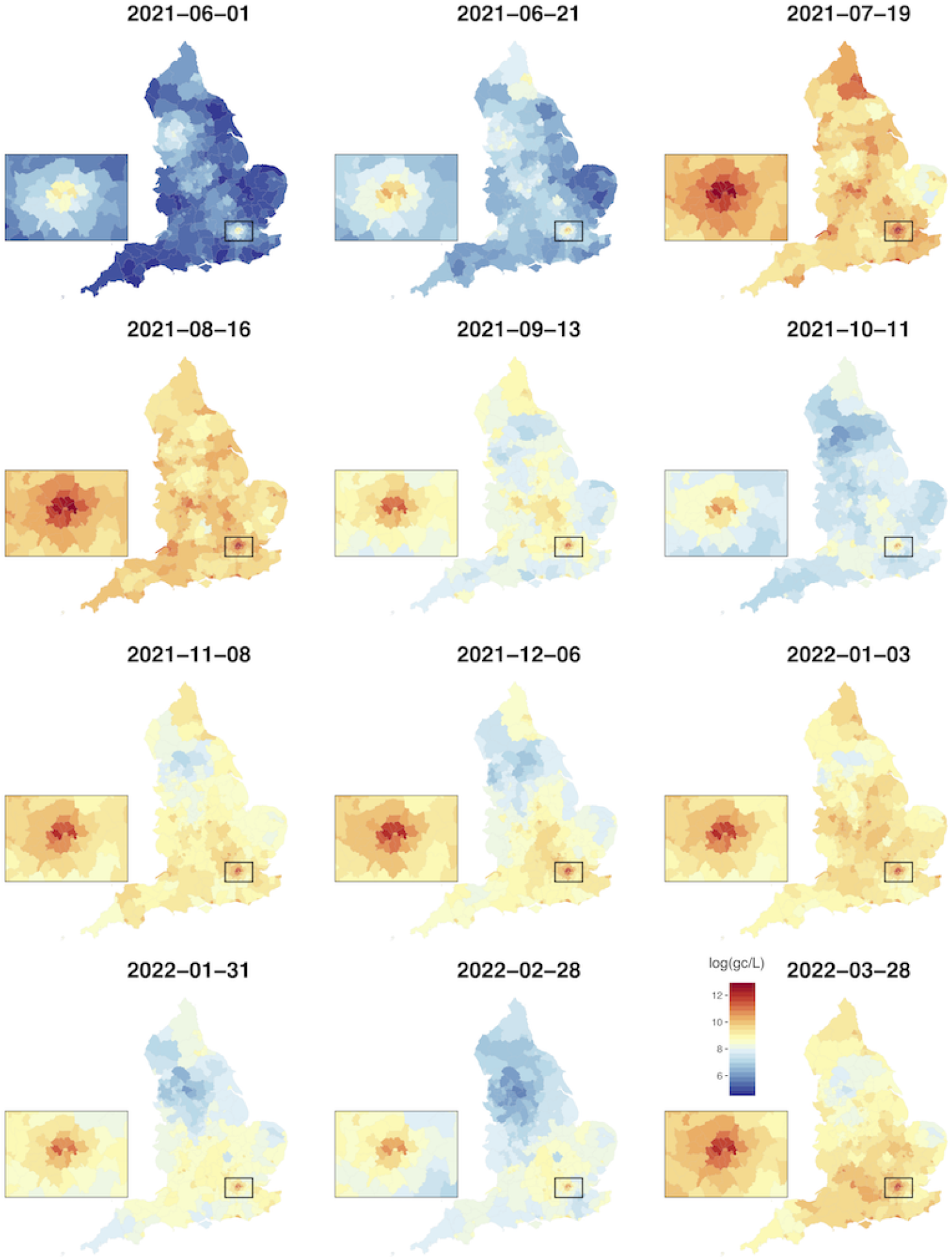
Spatio-temporal log-concentration. The figure shows the posterior predicted mean of the weekly concentration at LTLA level over June 2021 to March 2022. The dates shown are the Mondays of the weeks (apart from Tuesday, June 1, 2021). Red colors identify higher values (see legend in the bottom right plot). The insets visualise Greater London.

A similar pattern of gradual spatial spread can be seen between December 2021 and January 2022 and then between February and March 2022. For both periods, rises in concentration started principally in the south of England but then reached most parts of the country within the following month (Figure 4, second and third plots on the third and fourth rows). The extent of the spatio-temporal dynamics is quantified by the parameters of the spacetime interaction component, *z*_*it*_ in Eq. (2): the temporal AR1 coefficient, *ρ*, and the range parameter in the Matèrn covariance function, *r*, are estimated as 0.858 (0.822 - 0.899) and 59.53 km (52.82 - 68.78), respectively, revealing a fully spatio-temporal correlation structure. To visualise these rich model outputs, we have created a dynamic and interactive dashboard, accessible at https://b-rowlingson.gitlab.io/wwatlas/, that allows users to interrogate levels and changes in viral concentration at both local and national scales. The dashboard also features the weekly maps highlighting areas with sustained increases in concentration over consecutive weeks, using the rule described in Section 2.5.

### 3.3. Covariate effects

While the main aim of this analysis is to predict concentration at specific locations, covariate effects (Table 1) are also of interest. Population density is strongly associated with viral concentration in wastewater. The posterior mean of the corresponding regression coefficient is 0.278 (95% credible interval: 0.136 - 0.420). This corresponds to the amount of increase in log viral concentration when population density increases by about 1600 people per km^2^. Similarly, the percentage of genome coverage at national level is strongly associated with wastewater viral concentration, with a posterior mean of 0.518 (95% CI: 0.140 - 0.895), suggestive of a greater number of viral RNA fragments present in collected samples when the sample covers more of the SARS-CoV-2 genome. For the remaining covariates there is weaker evidence of an association with viral concentrations as their 95% credible intervals include 0 (see Table 1).

**Table 1:** Posterior estimates of covariate effects (one standard deviation change) and of parameters associated with the spatio-temporal random effects.

|  | **Posterior mean**<br>(95% credible interval) |
| --- | --- |
| **Covariates** |  |
| IMD | -0.032 (-0.139, 0.075) |
| BAME proportion | -0.056 (-0.196, 0.084) |
| Population density | 0.278 (0.136, 0.420) |
| % population aged below 16 | 0.057 (-0.083, 0.197) |
| % population aged over 75 | -0.052 (-0.201, 0.097) |
| Industrial fraction | -0.038 (-0.138, 0.062) |
| Genome coverage | 0.518 (0.140, 0.895) |
| SNP number | -0.038 (-0.273, 0.197) |
| **Hyperparameters** |  |
| $\sigma_y^2$, residual variance | 0.865 (0.835, 0.898) |
| $\sigma_g^2$, variance of the regional random effect | 0.027 (0.002, 0.051) |
| $\sigma_u^2$, variance of the spatial random effect | 0.388 (0.299, 0.494) |
| $\sigma_v^2$, variance of the temporal random effect | 0.253 (0.190, 0.344) |
| $r$, correlation range (km) in the Matérn covariance | 59.53 (52.82, 68.78) |
| $\sigma_z^2$, variance in the Matérn covariance | 0.509 (0.444, 0.595) |
| $\rho$, temporal AR1 coefficient | 0.858 (0.822, 0.899) |

### 3.4. Cross-validation

Cross-validation of predicted log-concentrations gives an overall mean bias, averaged across all sites and all time points, of 0.0093 log(gc/L) and the standard deviation of all biases is 1.27. No individual week within the observation period shows a particularly large mean bias (Figure 2 in supplementary material). The site-specific mean bias, averaged over the weeks with no missing data at each cross-validation site, ranges from -1.70 (Wycombe) to 3.48 (Burton on Trent). The variation in prediction quality across all sites does not appear to be associated with how close a cross-validation site is to an in-sample site (Figure 1 in supplementary material). The overall 95% coverage rate, representing the percentage of observed values falling within the corresponding 95% predictive interval, is 94.1%, which is close to the 95% nominal value thus indicating that our method provides reliable predictions. Table 1 in supplementary material provides further details on the model’s predictive performance. Performance of the model is not systematically different across the 10 cross-validation folds (Figure 3 in Supplementary material).

### 3.5. Wastewater concentration and COVID-19 debiased prevalence

We illustrate the correspondence between the predicted wastewater concentration and COVID-19 debiased prevalence over the period from 1 June, 2021 to 27 March, 2022. Debiased prevalence was estimated combining testing data and the REal-time Assessment of Community Transmission (REACT) randomised survey (Riley et al., 2020) using the method proposed by Nicholson et al. (2022); for more details on this method, see Supplementary Material, Section B.1). The prevalence estimates were made weekly for all LTLAs in England apart from City of London and Isles of Scilly. Specifically, Figure 5 maps the posterior probability of detecting an increase as described in Section 2.5 for the predicted wastewater concentration and for the estimated debiased prevalence over two specific periods, from 1 June, 2021 to 4 July, 2021 and from 4 October, 2021 to 7 November, 2021. We use a biscale legend where the green colours represent agreement between the two metrics. The first period coincides with a sharp increase in prevalence and we observe a good agreement between the two metrics, with most of the LTLAs moving from pale green (both metrics having posterior probability <0.8 of an increase over the previous 2 weeks) to dark green (both metrics having posterior probability >0.8 of an increase over the same period). Prevalence during the second period from 4 October, 2021 to 7 November, 2021 remained relatively stable, a pattern that we can also observe in the viral concentration estimates, thus giving rise to the almost completely pale green maps on the second row of Figure 5. Over the entire period from 1 June, 2021 to 27 March, 2022, 79.7% of the LTLA-three-weekly comparisons show concordance between wastewater viral concentration and COVID-19 prevalence with both metrics having >0.8 in probability of a rise over the previous two weeks or both having <0.8 in probability of a rise over the same time period. While the two metrics are in good agreement, we found that the relationship between viral concentration in wastewater and COVID-19 prevalence is complex, nonlinear and varying over both space and time (see Supplementary Material, Figures 4-5).

**Figure 5:**
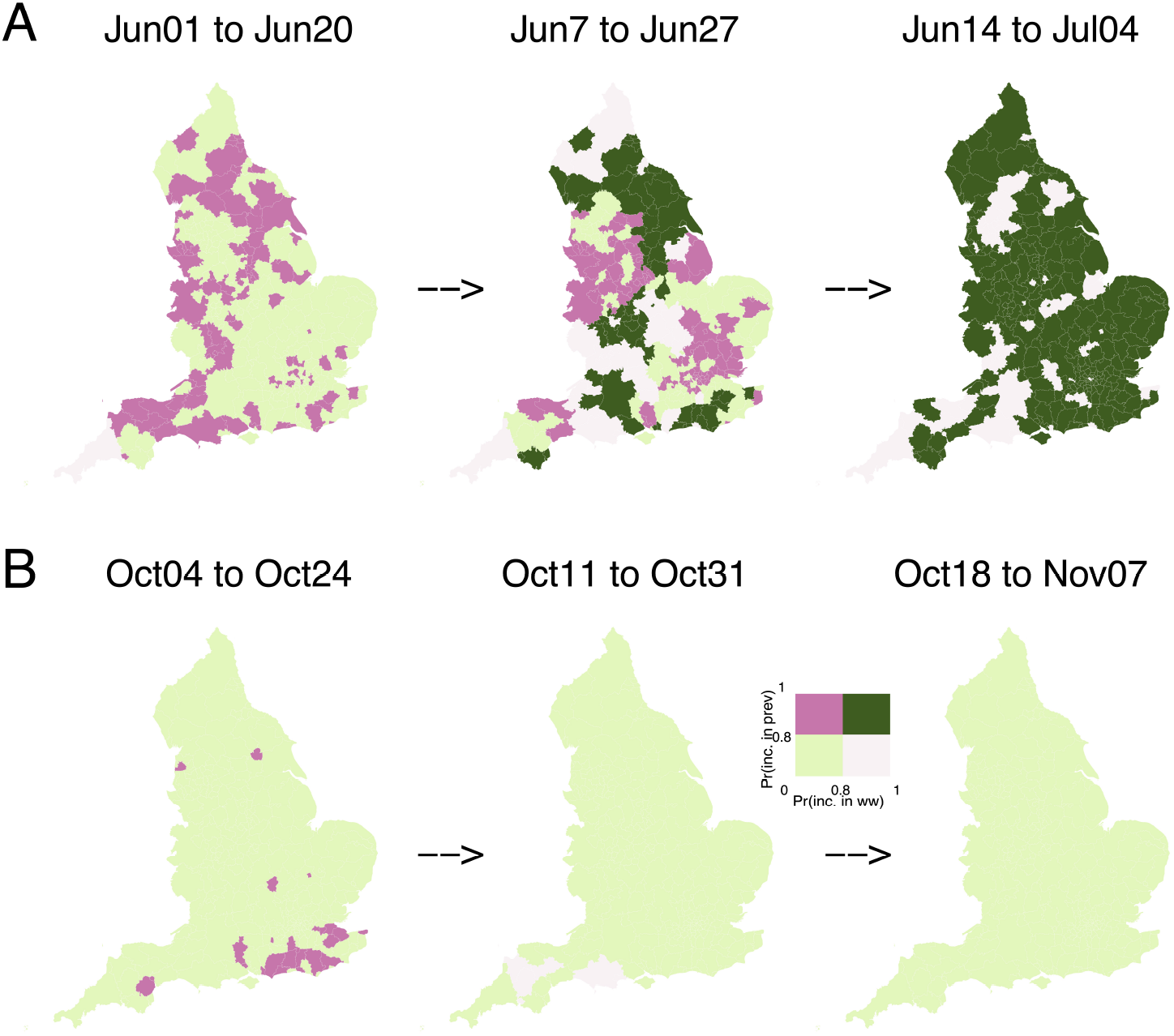
Correspondence between changes in wastewater viral concentration and changes in debiased prevalence. We show two periods: A. June 1 - July 4 2021 and B. October 4 - November 7 2021. Each LTLA is colour coded such that a dark green colour represents high probability (>0.8) of rise in both viral concentration and prevalence. Pale green indicates an LTLA at which neither viral concentration nor prevalence exhibited high probability of rise. Dark pink and light pink indicate, respectively, LTLAs where rise was detected in prevalence but not in viral concentration and those where rise was detected in viral concentration but not in prevalence.

## 4. Discussion

In this paper we have proposed a geostatistical approach to model wastewater viral concentration at STW catchment areas as a function of covariates, while accounting for residual spatio-temporal correlation. To the best of our knowledge this is the first study to go beyond catchment-level estimates by predicting viral concentrations on a spatially resolved domain, which can then be aggregated to any required spatial resolution to inform public health decisions in near-real-time. Specifically, we inferred the posterior predictive distribution on the population-weighted centroids of the 32,844 LSOAs across England and then aggregated these predictions to LTLA level, a geographical scale more relevant for public health policy-setting. Additionally, our approach uses probability statements to detect areas characterised by sustained increases in concentration over a specified period, providing a tool for early warning of local outbreaks.

This study provides a necessary foundation for investigating the link between wastewater and COVID-19 prevalence at any desired spatial resolution. A previous study (Morvan et al., 2022) considered the relationship between wastewater concentration and COVID-19 prevalence using the ONS-CIS survey and testing data on 45 STW catchment areas over the period July 2020 to March 2021. Specifying a spatio-temporal Bayesian model, they reported an overall good correspondence between the wastewater concentration and prevalence, concluding that wastewater can be used to provide reliable estimates of COVID-19 infections. Our approach similarly demonstrates the potential use of wastewater to track the space-time evolution of prevalence, but also provides a comparison at LTLA level across England, while Morvan et al. (2022) focused only on the ONS-CIS sub-regions intersecting with the catchment areas.

In line with Faraway et al. (2022) our results suggest that the spatiotemporal relationship between wastewater and prevalence is dynamic, complex and potentially nonlinear. For example, Spearman rank correlation between contemporaneous weekly LTLA-level predicted values of viral concentration and debiased estimates of COVID-19 prevalence reported by Nicholson et al. (2022), although predominantly positive, varied between about −0.25 and +0.75 over the whole of the study-period; see Figure 4 in Supplementary Material. The variability in the correlation could be driven by shedding still present in the vaccinated population (Nattino et al., 2022). Vaccination can also influence age-distribution of the disease, which in turn is related to shedding in wastewater (Sanjuán and Domingo-Calap, 2021). Additionally, external variables such as meteorology can play a role in diluting viral concentration (Foladori et al., 2021). All these aspects need to be accounted for in a principled way, calling for future research on modelling the relationship between wastewater and prevalence. Nevertheless, we have demonstrated a high concordance between local week-on-week *increases* in wastewater concentration and debiased prevalence (Figure 5), suggesting that the difficulties of calibrating current values of these two metrics need not prevent the use of wastewater-based epidemiology as a cost-efficient method for producing early warnings of local outbreaks that could be followed up by additional, more intensive sampling and/or focused public health action. In this study we have used weekly data, but recognise that daily data may be better suited to assessment of leads or lags between the signals from wastewater and from traditional epidemiological metrics. However, this would present additional modelling challenges to account for potential weekly periodicity, the irregular pattern of wastewater sampling days leading to missing values (Safford et al., 2022) and the relatively greater variability in the lag between the times of infection and detection.

To conclude, in this paper we present a geostatistical framework that can be used for predicting wastewater concentrations over a continuous spatial domain. Our predictions are based on sampling from the predictive distribution of the complete spatio-temporal surface of concentrations over the area of interest. They can therefore be transformed directly into samples from the predictive distribution of any required summary of this surface. For example, they can be aggregated to any given set of spatial units that is most relevant for public health policy, or can be used to detect locations that, with high probability, show a sustained increase over a period of time. Also, the probabilistic basis of the predictions delivers measures of predictive uncertainty that can inform adaptive sampling strategies, for example by directing additional sampling effort at areas of high uncertainty (Srinivas et al., 2021). Our approach should be seen as a starting point for the development of a cost-effective public health surveillance framework in which wastewater-based epidemiology can play a valuable role. However, we stress that any such framework should be extended to include other data sources, such as randomised surveys and testing data, to enable quantitatively accurate monitoring of disease evolution across a population.

## Data Availability

All data produced are available online at https://github.com/gqlNU/publicWW/

https://b-rowlingson.gitlab.io/wwatlas/

https://github.com/gqlNU/publicWW/

## 5. Author contributions

**Guangquan Li**: Conceptualization, Methodology, Data analysis, Writing-Original draft preparation.

**Hubert Denise**: Data curation.

**Peter Diggle**: Conceptualization, Funding, Supervision, Writing-Reviewing and Editing.

**Jasmine Grimsley**: Conceptualization.

**Chris Holmes**: Conceptualization, Funding.

**Daniel James**: Conceptualization, Writing-Reviewing and Editing.

**Radka Jersakova**: Visualization, Software, Writing-Reviewing and Editing.

**Callum Mole**: Visualization, Software.

**George Nicholson**: Data curation, Writing-Reviewing and Editing.

**Camila Rangel Smith**: Visualization, Software.

**Sylvia Richardson**: Conceptualization, Funding, Supervision.

**William Rowe**: Data curation.

**Barry Rowlingson**: Visualisation, Software, Writing-Reviewing and Editing.

**Fatemeh Torabi**: Writing-Reviewing and Editing.

**Matthew Wade**: Conceptualisation, Writing-Reviewing and Editing.

**Marta Blangiardo**: Conceptualization, Methodology, Supervision, Writing-Original draft preparation.

## 6. Acknowledgments

We would like to thank Chris Lilley (UKHSA) and Brieuc Lehmann (UCL) for providing some of the data for this work. We also acknowledge the Environmental Monitoring for Health Protection programme and its partners, including The Environment Agency.

