## Supplementary Material for "A spatio-temporal framework for modelling wastewater concentration during the COVID-19 pandemic"

Guangquan Li et al.

Supplementary Material

#### Contents

**A Cross-validation** **1**

**B Comparison with debiased prevalence** **4**
B.1 Debiased prevalence estimation . . . . . . . . . . . . . . . . . . . . . . . . . . . . . . . . 4
B.2 Correlation between wastewater concentration and debiased prevalence . . . . . . . . . . . 4

**C References** **7**

#### A Cross-validation

Table 1 summarises the mean bias, mean absolute bias, root mean square error and the 95% coverage by region across all the 303 sewage treatment works (STWs) included in the cross-validation. Figure 1 shows no clear relationship between the site-level mean absolute bias and the distance of each cross-validated site to its closest in-sample STW. Figure 2 shows that no one week has a large mean bias across the study period and Figure 3 shows no systematic difference in bias over the 10 cross-validation folds.

| | MB | MAB | RMSE | 95% coverage |
|---|---|---|---|---|
| East Midlands | -0.02 | 0.99 | 1.25 | 0.95 |
| East of England | 0.03 | 1.04 | 1.34 | 0.93 |
| London | -0.04 | 0.91 | 1.12 | 0.97 |
| North East | -0.07 | 0.77 | 0.99 | 0.98 |
| North West | 0.04 | 1.02 | 1.31 | 0.92 |
| South East | -0.02 | 0.98 | 1.27 | 0.94 |
| South West | -0.05 | 0.91 | 1.14 | 0.97 |
| West Midlands | 0.05 | 1.06 | 1.38 | 0.93 |
| Yorkshire and The Humber | 0.14 | 1.07 | 1.36 | 0.92 |

Table 1: Summary of the mean bias (MB), mean absolute bias (MAB), root mean square error (RMSE) and the 95% coverage by the nine English regions. Bias is defined as the difference between the predicted and the observed values. Mean is taken across all STWs within a region over the period between June 1, 2021 and March 30, 2022.

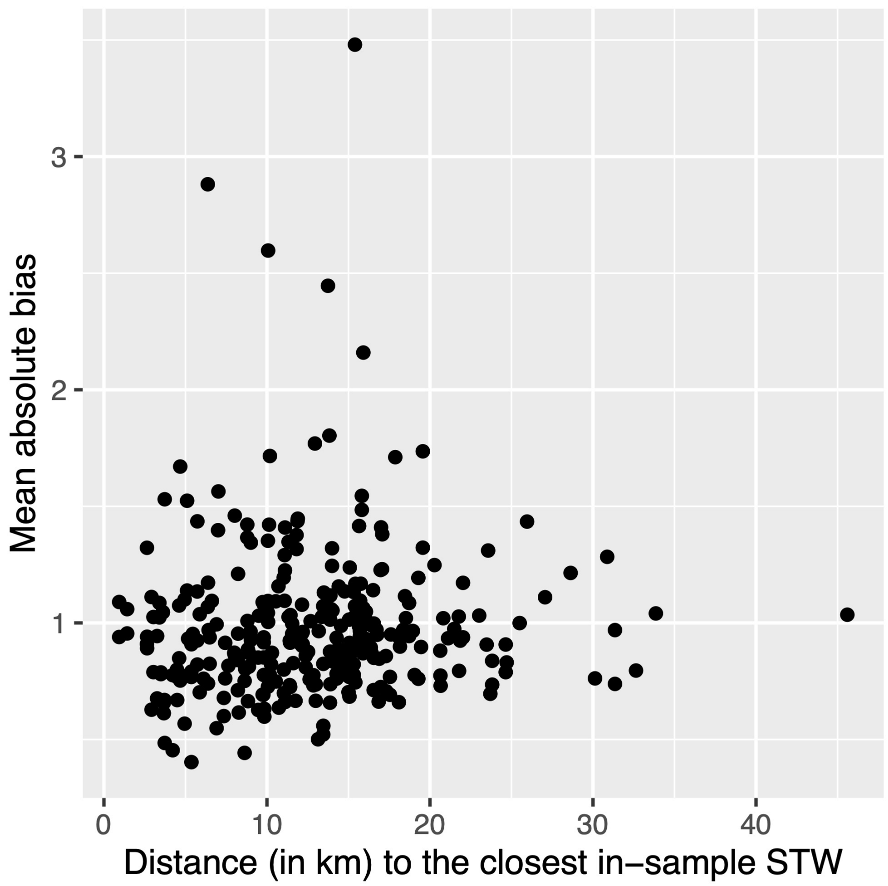


Figure 1: Relationship between the mean absolute bias and the distance to the closest in-sample STW site.

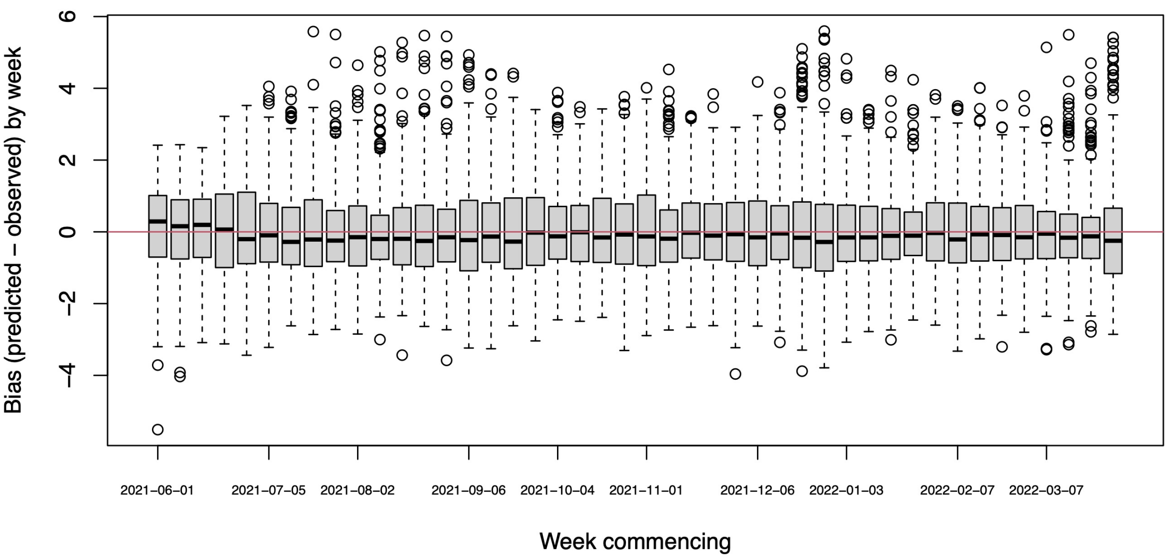


Figure 2: A boxplot examining variation in bias across the 44 weeks in the study period.

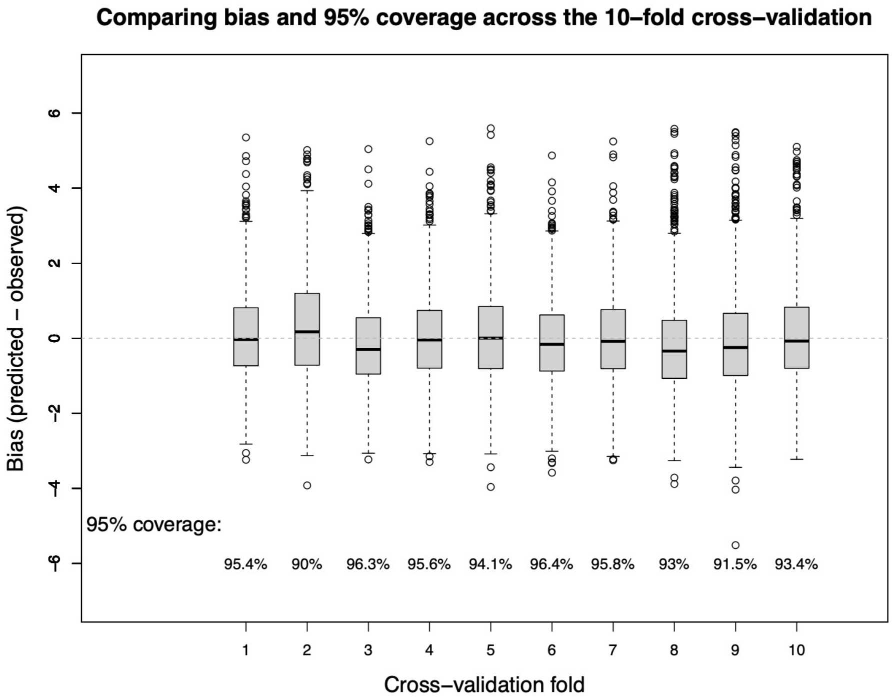


Figure 3: A boxplot summarising the biases over the 10-fold cross validation. Also reported are the 95% coverage by cross-validation run.

### B Comparison with debiased prevalence

#### B.1 Debiased prevalence estimation

We use the output of the modelling framework of Nicholson et al.,(2022), to estimate weekly COVID-19 prevalence at LTLA covering the study period. Test positivity rates in Pillar 2 data are strongly biased upwards relative to the population prevalence proportion, as the testing is directed at the higher risk population (e.g. symptomatics, frontline workers). We showed in Nicholson et al.,(2022) how careful modelling of the ascertainment process allows us to estimate prevalence accurately, and with good precision, even at a fine-scale level such as LTLA.

There we introduced the following causal model for the observation of positive of total positive targeted (e.g. Pillar 2) tests:

$$\begin{aligned}\mathbb{P}(n \text{ of } N \mid, \delta, \nu) &= \text{Binomial}\left(n \mid I, \mathbb{P}(\text{Tested} \mid \text{Infected})\right) \\ &\times \text{Binomial}\left(N-n \mid M-I, \mathbb{P}(\text{Tested} \mid \text{Not Infected})\right) \end{aligned} \tag{1}$$

where $\delta$ and $\nu$ parameterise (on log odds scale) the binomial success probabilities $\mathbb{P}(\text{Tested} \mid \text{Infected})$ and $\mathbb{P}(\text{Tested} \mid \text{Not Infected})$:

$$\delta := \log\left(\frac{\text{Odds}(\text{Tested} \mid \text{Infected})}{\text{Odds}(\text{Tested} \mid \text{Not Infected})}\right) \tag{2}$$

$$\nu := \log \text{Odds}(\text{Tested} \mid \text{Not Infected}) \, . \tag{3}$$

The parameter requiring careful treatment is $\delta$, i.e. the log odds ratio of being tested in the infected versus the non-infected subpopulations. This ascertainment parameter $\delta$ is estimable by combining accurate prevalence information from a randomised surveillance study (REACT) with the targeted testing data from Pillar 2. (The other parameter, $\nu$, is directly estimable from the targeted data, with $\hat{\nu} := \text{logit}[(N-n)/M]$ acting as a precise estimator with little bias when prevalence is low.) For further details of the prevalence debiasing model, including validation against local randomized surveillance data, please see Nicholson et al.(2022).

#### B.2 Correlation between wastewater concentration and debiased prevalence

Figure 4 presents correlation between viral concentration and COVID-19 debiased prevalence over the period from June 1, 2021 to March 27, 2022. Over time (Fig. 4A), levels of viral concentration in wastewater were highly positively correlated with debiased prevalence between June and August 2021 and between mid-November and end of March 2022, suggesting that an LTLA with high viral concentration tends to show high COVID-19 prevalence within the same week. The levels of the two metrics, however, appear to be unrelated and even negatively correlated between mid-August and early November 2021. Fig. 4B shows the spatial variability of the correlation between viral concentration and prevalence. Of the 307 LTLAs, 222 LTLAs have posterior probabilities exceeding 0.95 that the weekly viral concentrations are positively correlated with the weekly debiased prevalence (Fig. 4C). We used Spearman's rank correlation due to the nonlinear correlation patterns between viral concentration and debiased prevalence (Fig. 5).

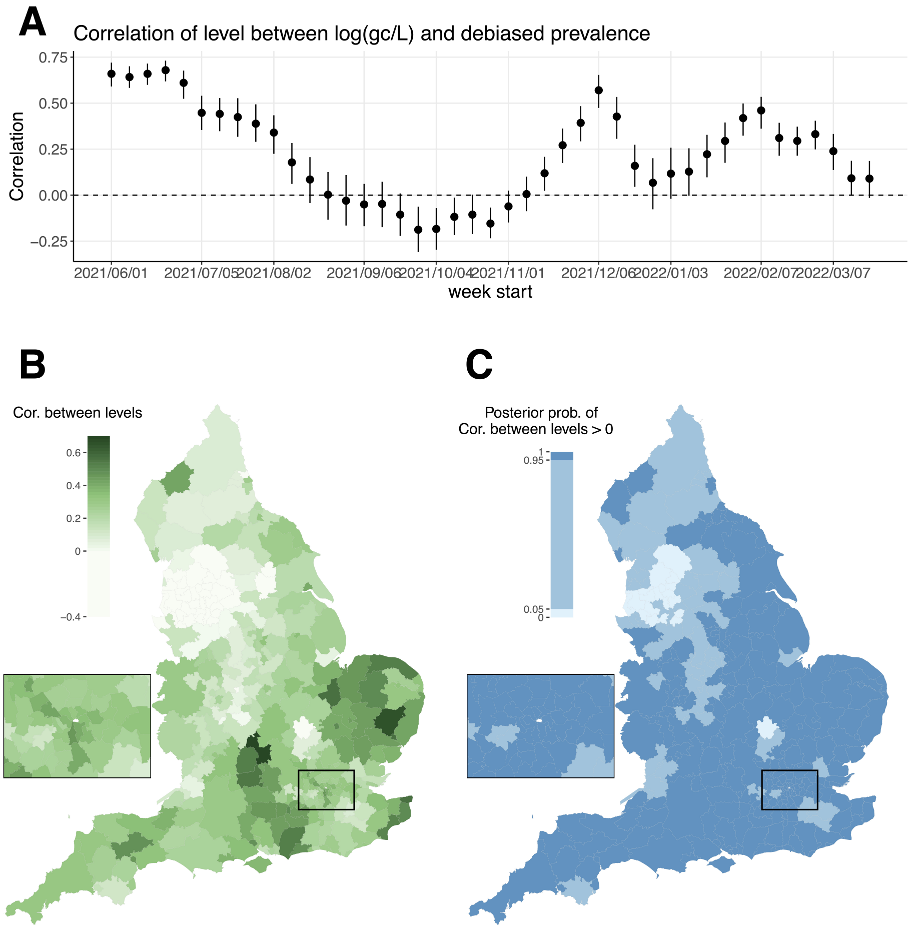


Figure 4: Correlation between viral concentration and debiased prevalence levels over time and space. Panel A: the posterior mean correlation across all 307 LTLAs by given week with the 95% credible interval. Panel B: the posterior means of the correlation by LTLA across all weeks. A dark green colour indicates high debiased prevalence tends to be associated with high viral concentration within the same week. Panel C: the posterior probability of the correlation being greater than 0. Debiased prevalence is not available for two LTLAs, City of London (the white polygon in the inset map) and Isles of Scilly (a group of islands off the southwestern tip of England; see Fig. 3B).

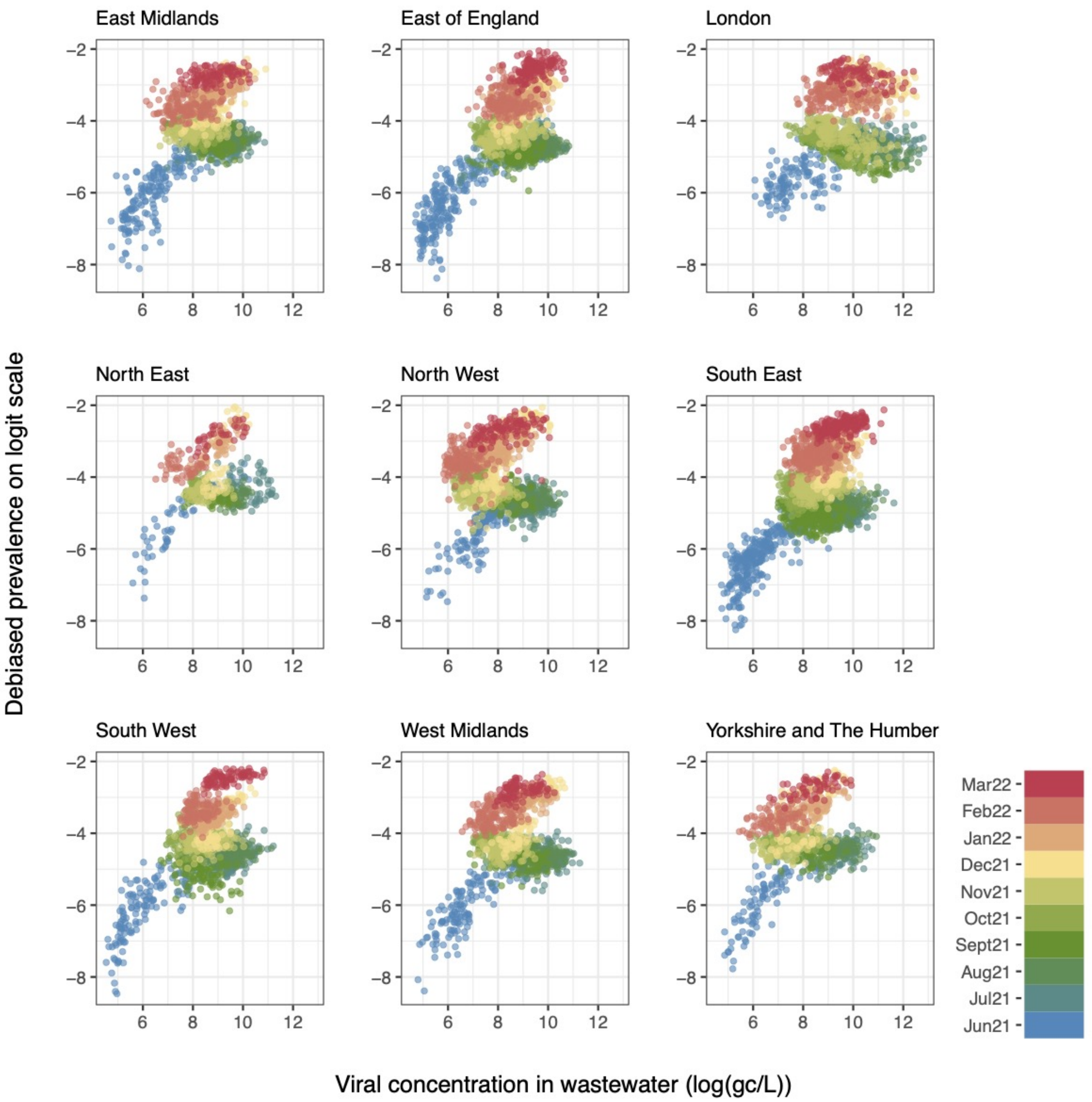


Figure 5: Scatterplots between viral concentration and debiased prevalence levels at the LTLA-weekly scale by region, showing the nonlinear feature of the wastewater-prevalence relationship. Points are colour-coded by month (see legend in the bottom right plot).
